# FDA approved Artificial Intelligence and Machine Learning (AI/ML)-Enabled Medical Devices: An updated landscape

**DOI:** 10.1101/2022.12.07.22283216

**Authors:** Geeta Joshi, Aditi Jain, Shalini Reddy Araveeti, Sabina Adhikari, Harshit Garg, Mukund Bhandari

**Affiliations:** Department of Urology, UTHealth San Antonio, TX; Department of Medical Education, UTHealth San Antonio, TX; Department of Obstetrics and Gynecology, UTHealth San Antonio, TX; New England College, NH; Department of Computer Science, Texas A&M University, College Station, TX; Greehey Children Cancer Research Center, UTHealth San Antonio, TX

**Keywords:** Machine Learning, Artificial Intelligence, FDA, Medical Devices, AI, ML, Clinical Trails

## Abstract

As artificial intelligence (AI) has been highly advancing in the last decade, machine learning (ML) enabled medical devices are increasingly used in healthcare. In this article, we performed comprehensive analysis of FDA approved Artificial Intelligence and Machine Learning (AI/ML)- Enabled Medical Devices and offer an in-depth analysis of clearance pathways, approval timeline, regulation type, medical specialty, decision type, recall history etc. We found a significant surge in approvals since 2018, with clear dominance of radiology specialty in the application of machine learning tools, attributed to the abundant data from routine clinical data. The study also reveals a reliance on the 510(k)-clearance pathway, emphasizing its basis on substantial equivalence and often bypassing the need for new clinical trials. Also, it notes an underrepresentation of pediatric-focused devices and trials, suggesting an opportunity for expansion in this demographic. Moreover, the geographical limitation of clinical trials, primarily within the United States, points to a need for more globally inclusive trails to encompass diverse patient demographics. This analysis not only maps the current landscape of AI/ML-Enabled Medical Devices but also pinpoints trends, potential gaps, and areas for future exploration, clinical trial practices, and regulatory approaches.

## Introduction

The term “Artificial Intelligence” (AI) was first introduced by John McCarthy in 1956 during a seminal conference for a summer research project[1]. AI is characterized as the ability of a computer program to perform a wide array of tasks that typically require human intelligence. These tasks include, but are not limited to, reasoning and learning. With the widespread integration of AI applications, the field has evolved, encompassing several significant subsets as depicted in Figure 1. However, it is critical to note that terminologies within this domain, specifically artificial intelligence, machine learning (ML), and deep learning (DL), have often been used interchangeably.

Machine learning, a subset of AI, involves systems that can assimilate knowledge from data over time and use this information to make predictions upon the introduction of new, unseen datasets. Another subset, Deep Learning (DL), employs sophisticated algorithms known as neural networks, which are designed to mimic the neural structures of the human brain[2]. These algorithms are widely applied in numerous fields, including but not limited to speech recognition, computer vision, drug discovery, and genomics[3]. The efficacy and robustness of these models are substantially enhanced as they are exposed to more comprehensive, diverse, and heterogeneous data, enabling the models to achieve greater accuracy and reliability in their predictive capabilities[4].

Machine learning has garnered widespread utilization in medical research, primarily due to its ability to extract pertinent clinical insights from the voluminous data generated within the healthcare sector daily[5] . There has been a notable trend in integrating machine learning models into medical devices[6] [7], reflecting the growing influence of computational technology in various aspects of healthcare and clinical diagnostics.

In the United States, adherence to regulatory standards is mandatory for the commercial operation of technologically advanced medical devices and tools. The Food and Drug Administration (FDA) regulates medical devices and approves their market entry through one of three distinct regulatory pathways: Premarket Notification 510(k) clearance[8], De Novo (DEN) review[9], or Premarket Approval (PMA) [10], each detailed in Supplementary Table 1. This rigorous process ensures that any machine learning-augmented medical device adheres to safety and efficacy standards before its clinical use, thereby safeguarding public health while facilitating medical advancements.

In this brief, we discuss the most recent and current landscape of AI/ML-enabled medical devices approved by the FDA in the United States, reflecting developments up to the latest update on October 19, 2023, which included the addition of 171 new medical devices.

## Methods

We compiled a list of FDA-approved AI/ML-enabled devices across medical disciplines and assembled corresponding information for each device using the FDA’s publicly available data, as well as information provided by the specific manufacturers in their public notifications.

The inclusion criteria for AI/ML-based medical devices in our research encompass the devices listed on the FDA’s webpage in its most recent update on October 19, 2023. Once the inclusion criteria were established, we used the unique submission number of each device and manually extracted all the important features of the AI/ML-enabled medical devices. These features include the date of approval, the number of days taken to obtain clearance, clearance type (approval path), regulation panel, decision type, the name of the manufacturing company that filed for clearance, the country where the manufacturing company is based, device name, device medical specialty, device type and recall history.

Additionally, wherever available, we also gathered clinical trial information such as study type, sampling method, age group of subjects, criteria for inclusion, the number of clinical trial locations, and the names of the countries where the clinical trials were conducted.

The list of AI/ML-enabled devices provided by the FDA is not exhaustive and comprehensive in its entirety because it does not include other AI/ML-based medical devices that have not yet been approved by the FDA, or medical devices that are not categorized as AI/ML-based. Furthermore, AI/ML-enabled devices that are accredited, certified, or approved by other regulatory agencies inside or outside the USA are beyond the scope of this paper. Also, there is no information on the number of AI/ML-enabled medical devices that have failed to obtain FDA approval. The FDA webpage states that most of the summaries available on the site are abridged for public access, provided by the application submitters, and may not necessarily contain comprehensive information.

Data mining, visualization, and statistical analysis were performed using the base package in R version 3.6.3 (R Foundation for Statistical Computing, Vienna, Austria).

## Results

### 1. Overall Trends

As of the latest update on October 19, 2023, the FDA has listed 691 approved AI/ML-Enabled Medical Devices, out of which 103 (about 15%) were approved in the year 2023 to date (Figure 2). The trend in the number of approvals per year began to gain significant attention since 2016, and there has been an increase in the number of approved devices each year since then. Based on the current data, the highest number of approvals in a year so far was 139 AI/ML-enabled FDA-approved medical devices in 2022 (Figure 2).

### 2. Medical Subspecialties

The FDA has approved AI/ML-Enabled medical devices across many medical subspecialties (Figure 3a). An important insight from this data is that the majority, 531 (about 77%), of FDA-approved AI/ML-Enabled Medical Devices belong to the radiology medical subspecialty. Radiology not only has the highest volume of submissions but has also witnessed the most consistent growth in AI/ML-enabled device submissions among all specialties. Radiology is followed by the cardiovascular subspecialty with 70 devices (about 10%), then Neurology with 20 devices, and Hematology with 15 devices (Figure 3a, Figure 3b). Other medical subspecialties include Gastroenterology & Urology with 11 devices, Ophthalmic with 9 devices, Anesthesiology and Clinical Chemistry with 6 devices each, Microbiology, and General and Plastic Surgery with 5 devices each, Pathology with 4 devices, General Hospital with 3 devices, Ear Nose & Throat (ENT) with 2 devices, and Dental, Immunology, Obstetrics and Gynecology, and Orthopedic with one device each. Two medical devices related to ENT were only listed in 2022 and 2023 (Figure 4).

### 3. Device Classification

The top device classes approved by the FDA include radiological image processing, radiological image processing software, computed tomography, computer-assisted triage and notification software, ultrasonic imaging, and nuclear magnetic resonance imaging, as indicated by the primary product code (Figure 4, Supplementary Table 2).

### 4. Clearance Pathway, Decision Type, and Recall Rate

As of October 19, 2023, 96.7% of the approved AI/ML-enabled devices were cleared via the 510(k) pathway. Only 2.9% were via De Novo approval, while Premarket Approval (PMA) was granted to about 0.4% of the total approved devices (Figure 5a). While the FDA’s primary public health mission is to ensure the safety and effectiveness of innovative devices, approval decisions are frequently based on substantial equivalence to their predecessors of a similar kind. The FDA approved approximately 97% of AI/ML-enabled devices based on Substantial Equivalence (SESE) criteria, with other decision types making up the minority (Figure 5b). Similarly, the number of approved devices reviewed by a 3rd party comprises only about 3% (Figure 5c). The FDA has issued recall notices for about 5% of AI/ML-enabled medical devices, citing various concerns.

### 5. Approval Wait Time

Approval wait time was calculated by counting the days passed between the date the FDA received the complete application and the decision date. The median wait time varied among different medical subspecialties and by decision year (Figure 6a and Figure 6b). Devices related to ENT, Radiology, and General and Plastic Surgery had shorter median wait times. Similarly, median wait times have averaged around 125 days since 2016.

### 6. Applicant Company

At the time of application, there were altogether 295 applicants for all the AI/ML-enabled devices seeking FDA approval. Among them, GE Healthcare, Siemens, and Canon are the top three applicants, filing the greatest number of applications for FDA clearance (Figure 7). It should be noted that this data does not take into account company mergers, acquisitions, device rebranding, or company renaming.

### 7. Leading Countries in AI/ML Enabled Medical Devices

Based on the applicant’s country of origin, the USA leads the list with a total of 357 (approximately 52%) FDA-approved AI/ML Enabled Medical Devices, followed by Israel with 56 devices (Figure 8a, Figure 8b). France, Japan, and China complete the top five, with 33, 29, and 26 FDA-approved medical devices, respectively. Figure 9 serves as the summary plot for all the devices.

### 8. Clinical Trials

Only 22 (about 3.2%) AI/ML-enabled medical devices approved by the FDA reported conducting clinical trials (Figure 10a). Among these 22 devices, interventional trials were conducted for 9 devices, and observational types of clinical trials were conducted for 13 devices (Figure 10b). The devices related to these clinical trials mainly belong to the cardiovascular and radiology medical specialties (Figure 10c). These clinical trials primarily enrolled adults/older adults as their subjects (Figure 10d). A medical device related to pediatric autism spectrum disorder diagnosis aid was specific to the child age group. All clinical trials were open to both males and females for enrollment, except one specifically for females, which was related to a breast imaging system. The number of clinical trials for a device ranged from a single clinical trial to a total of 4 clinical trials and from a single location to 20 different locations (Figure 10e). However, the data shows that clinical trials for 15 of these devices were conducted only in the USA, while clinical trials for 6 of these devices were conducted in two or more different countries (Figure 10f). The devices with clinical trial information are summarized in Figure 11.

## Discussion

In this study, we illustrated how the number of FDA-approved AI/ML-Enabled Medical Devices has seen a consistent upsurge in the United States since the inaugural approval of such a device in 1995. A noteworthy increasing trend in the authorization of these devices became prominent from 2018 onwards, accounting for approximately 90% of all approved devices. This substantial growth in the approval trajectory can be associated with the comprehensive evolution within the computing field, marked by advancements in hardware and software, the affordability of cloud storage solutions, the accessibility of expansive data sets, and significant investments from major corporations in fostering more sophisticated platforms[11] [12] [13] [14].

Radiology emerges as the predominant specialty for the application of AI/ML-Enabled Medical Devices. This prevalence is attributed to the routine prescription of radiological imaging in regular clinical assessments and successive patient consultations, thereby amassing substantial datasets[15]. These data reserves are invaluable, facilitating extensive research initiatives for scientists and providing a robust foundation for device manufacturers to innovate and enhance medical devices[16].

In terms of FDA approval, the majority of FDA approvals were approved through the 510(k)- clearance pathway, relying on the demonstration of substantial equivalence that circumvents the necessity for exhaustive clinical trials. Remarkably, only around 3% of the totality of approved devices have disclosed undertaking clinical trials, with a predominant focus on adult participants. This trend suggests a potential shortfall in the sphere of medical specialties targeting pediatric and young adult demographics, either indicating a challenge in the development of AI/ML-enabled instruments for these age brackets or a lack of comprehensive inclusivity within the clinical trials.

Moreover, the geographical span of these clinical trials exhibits a considerable limitation, confined within the US borders. Such a restriction could overshadow the diverse heterogeneity integral to global subject samples involved in clinical examinations[17]. Moving forward, there is a pressing need to develop AI/ML based medical devices for medical specialties that are lagging behind and also broaden the demographic and geographic spectrum of clinical trials to encompass a more representative global populace, including underrepresented communities, to address disparities.

## Conclusion

This article succinctly delineates the integration of artificial intelligence and machine learning in the medical devices, highlighting the pertinent FDA approval channels. Furthermore, we have discussed major insights and summarized the spectrum of FDA-approved AI/ML-Enabled Medical Devices utilizing the limited publicly accessible information available up to the present date. Our analysis serves as a beacon, elucidating the current landscape of AI/ML-Enabled Medical Devices, and prevailing trends, thereby contributing to a broader understanding.

## Data Availability

All data produced are available online at FDA webpage.

---

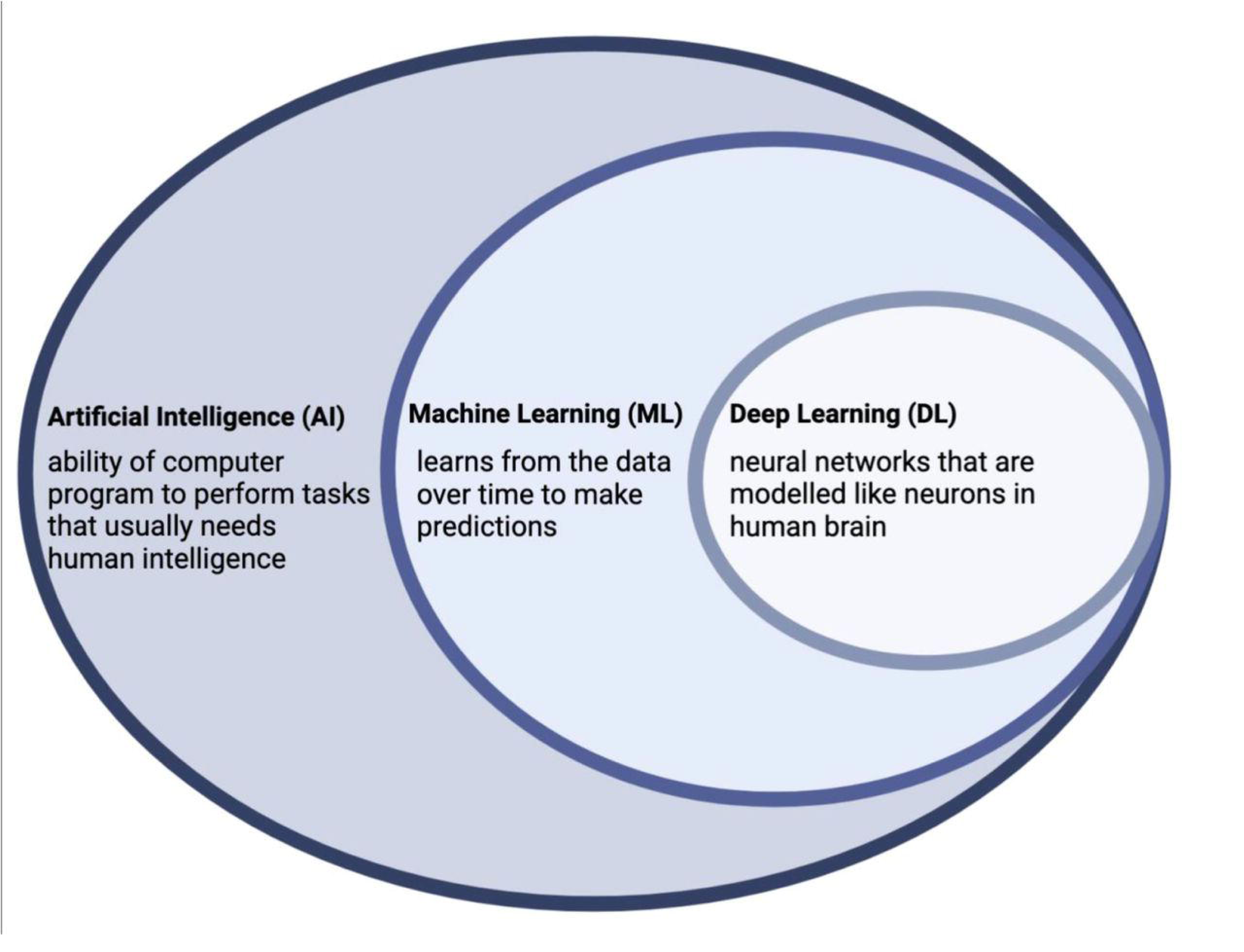

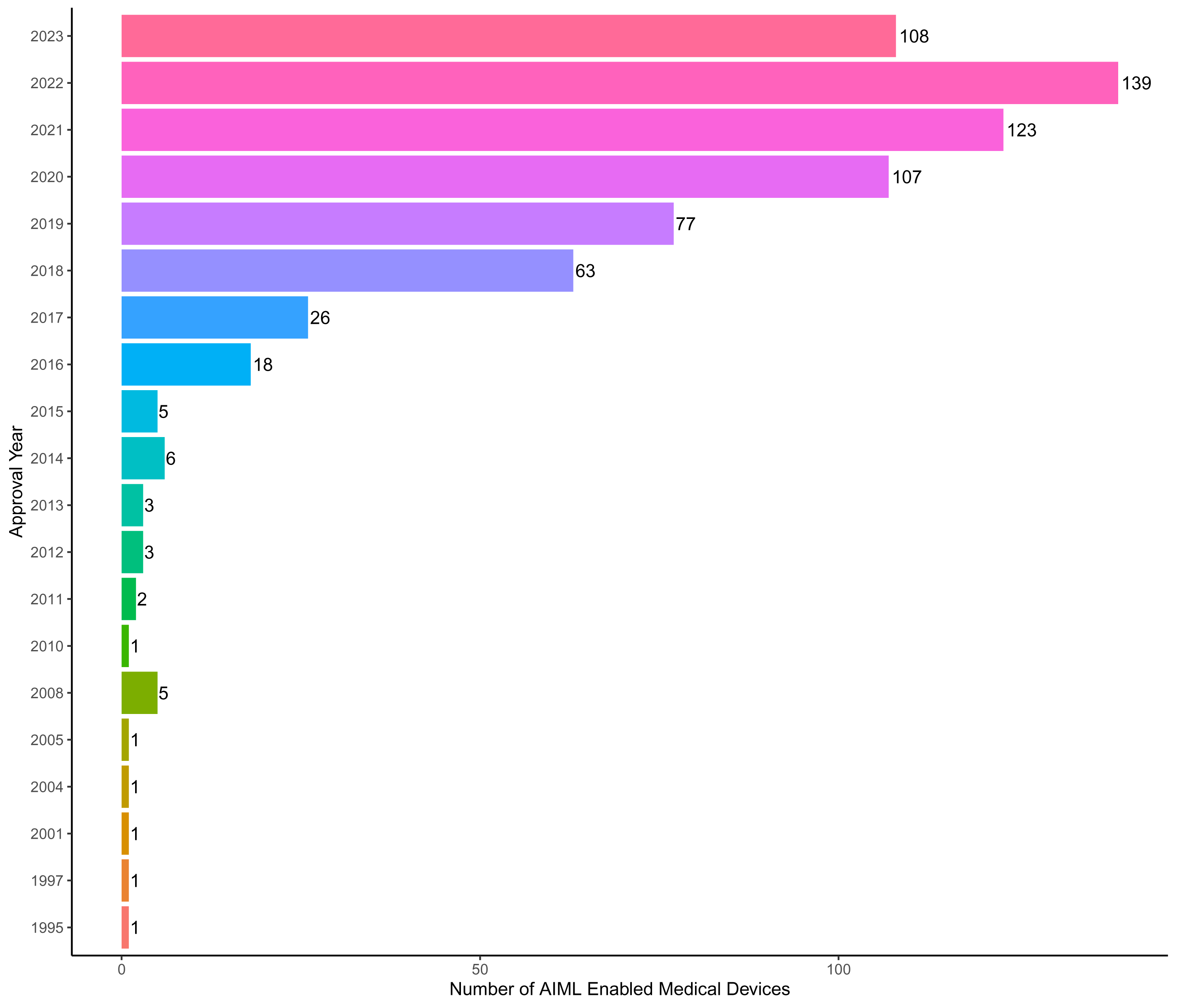

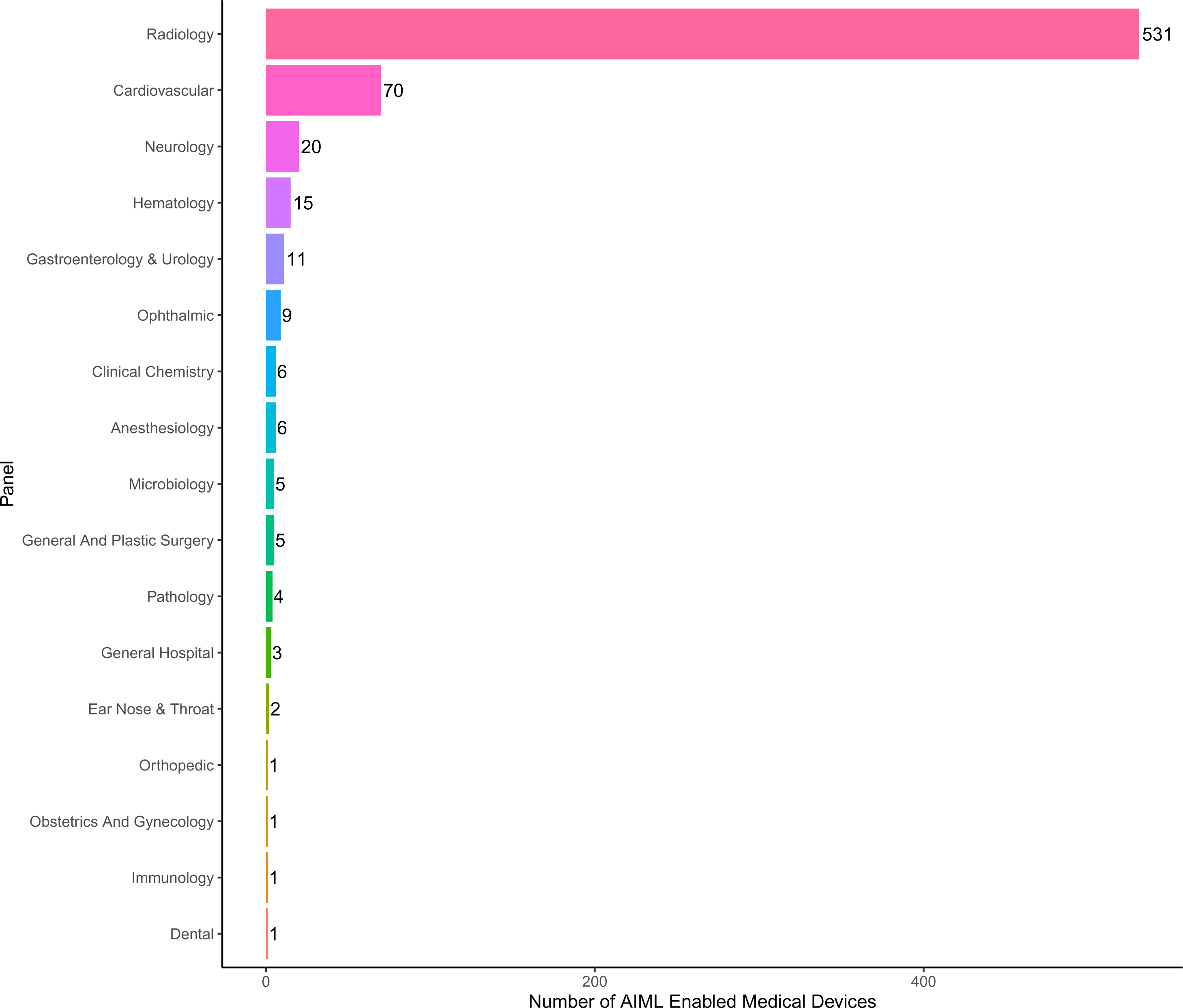

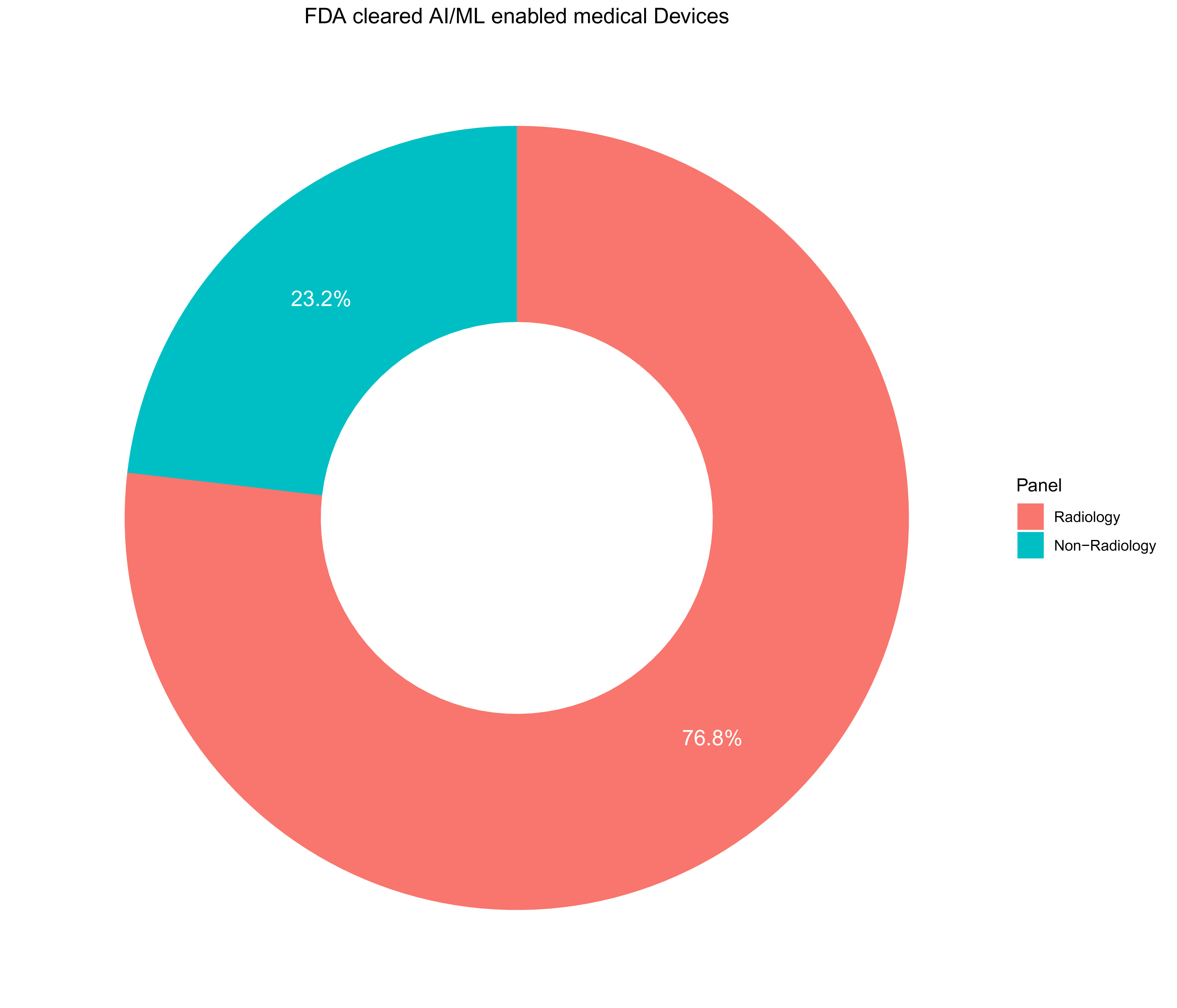

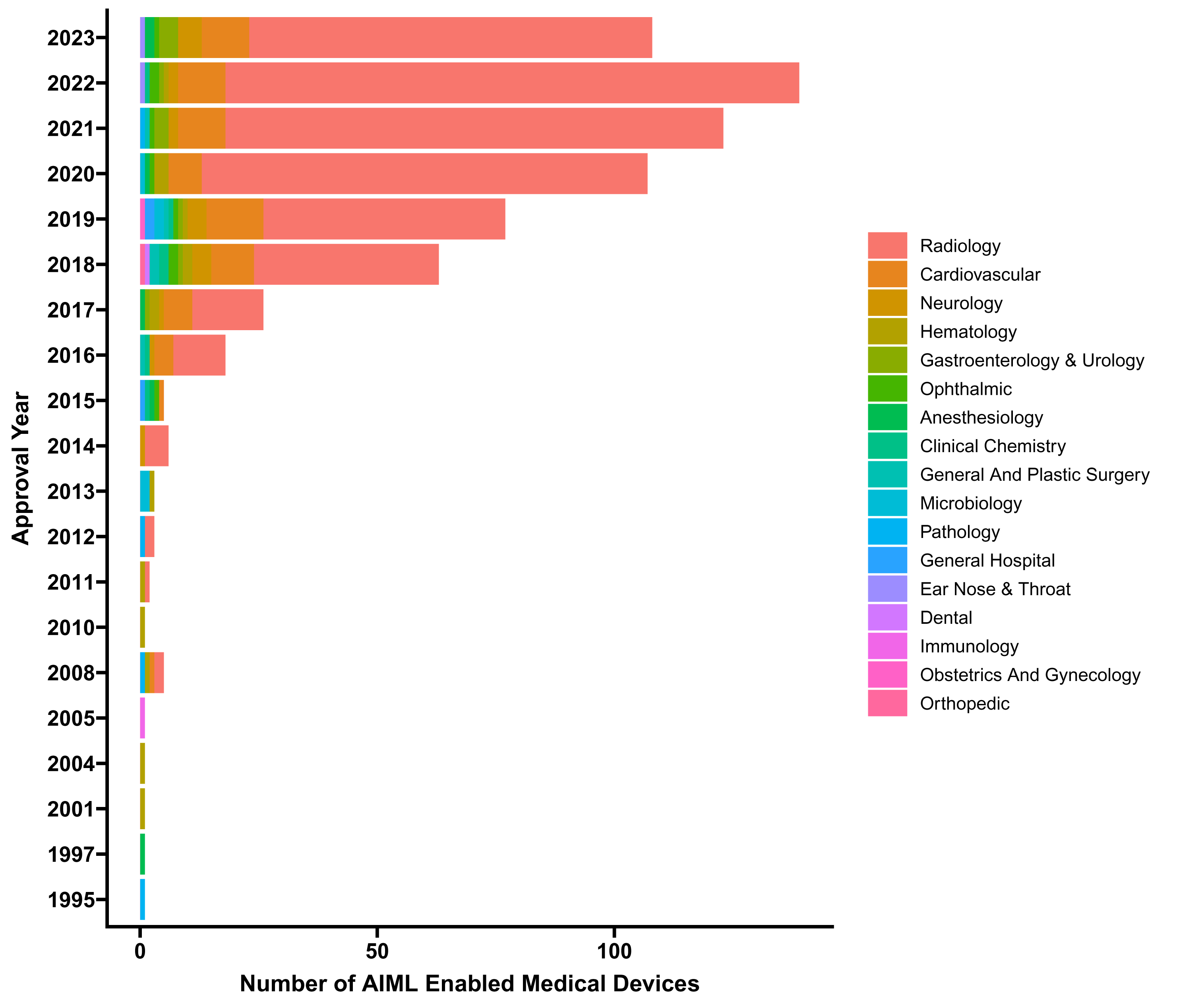

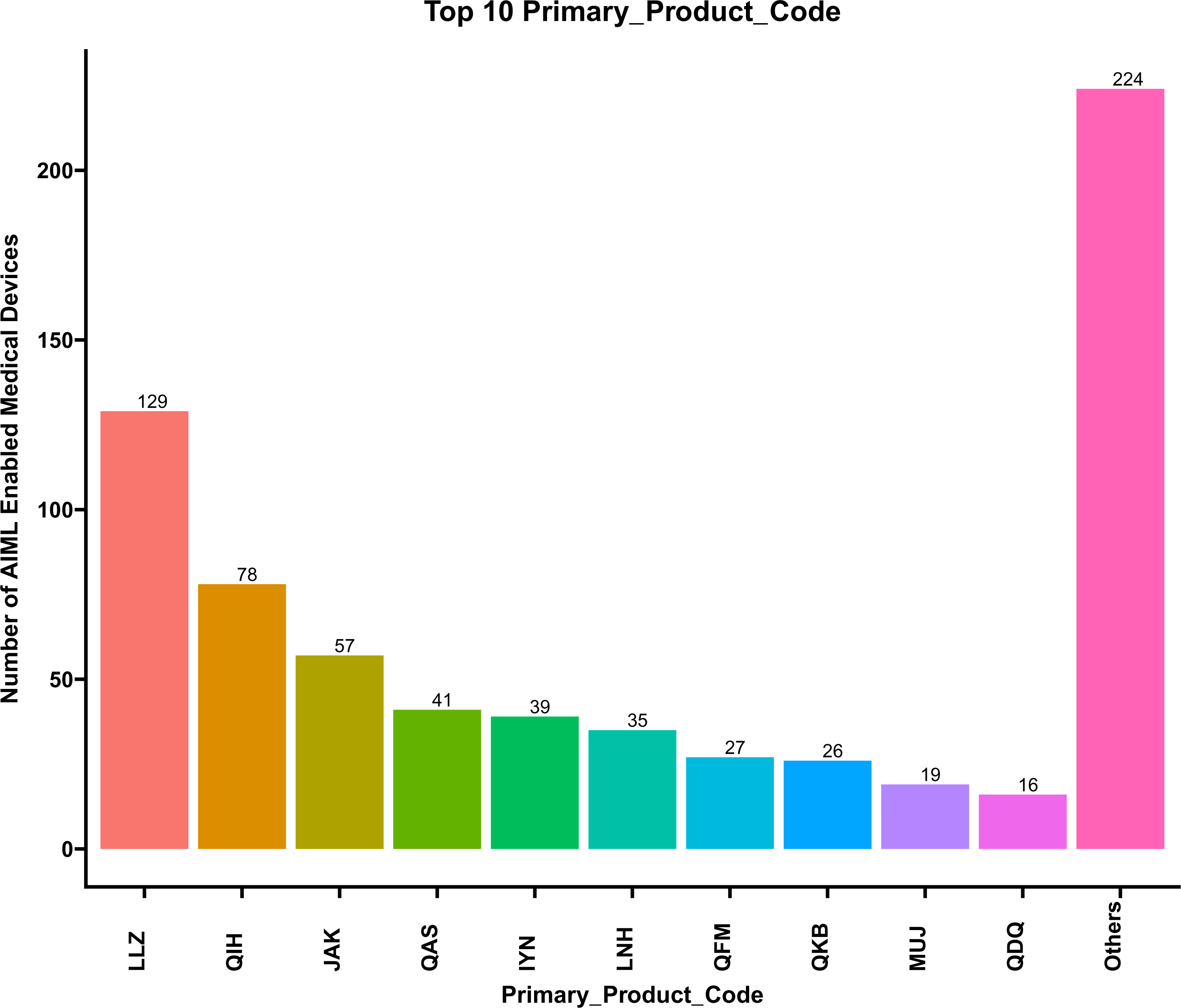

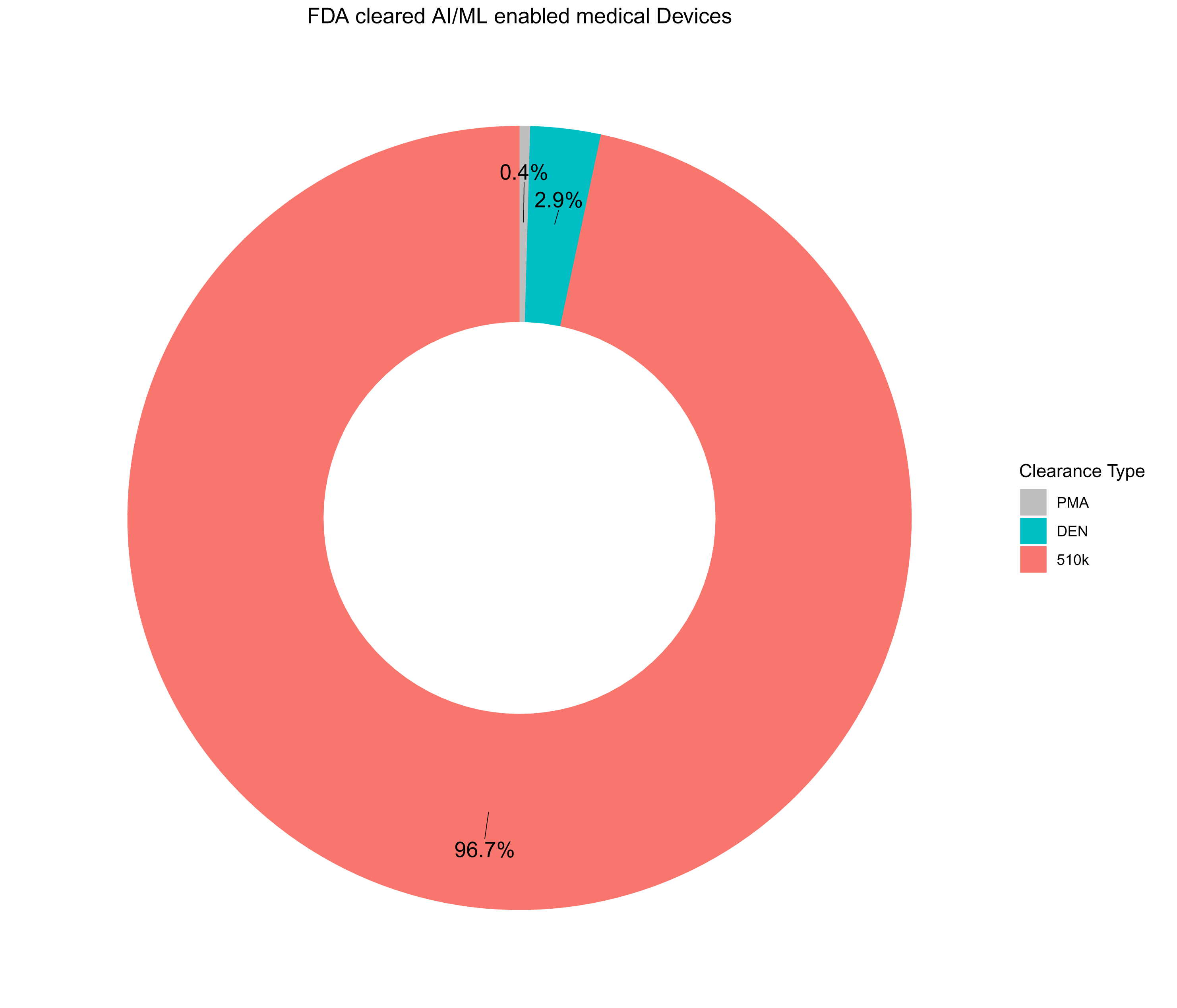

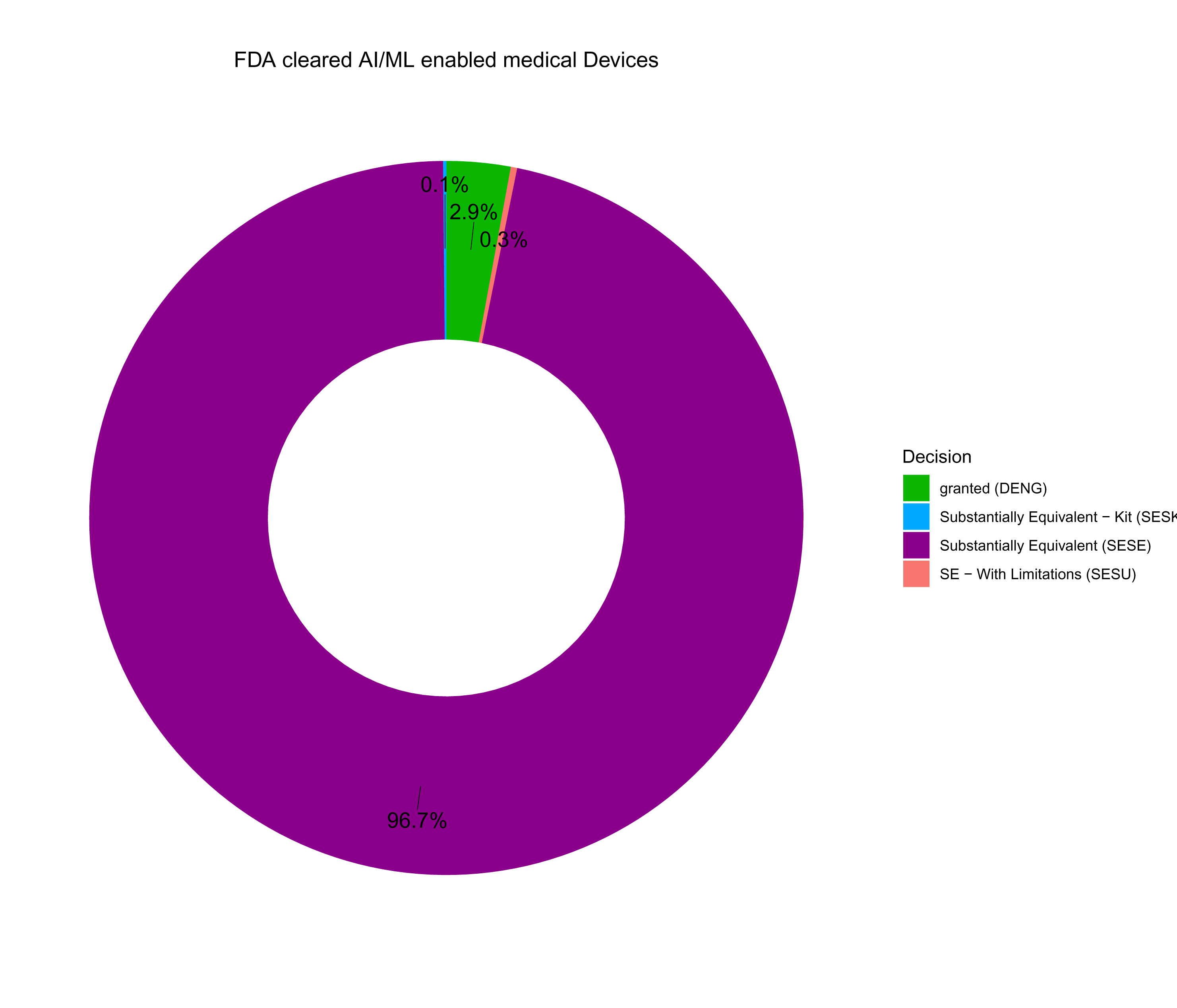

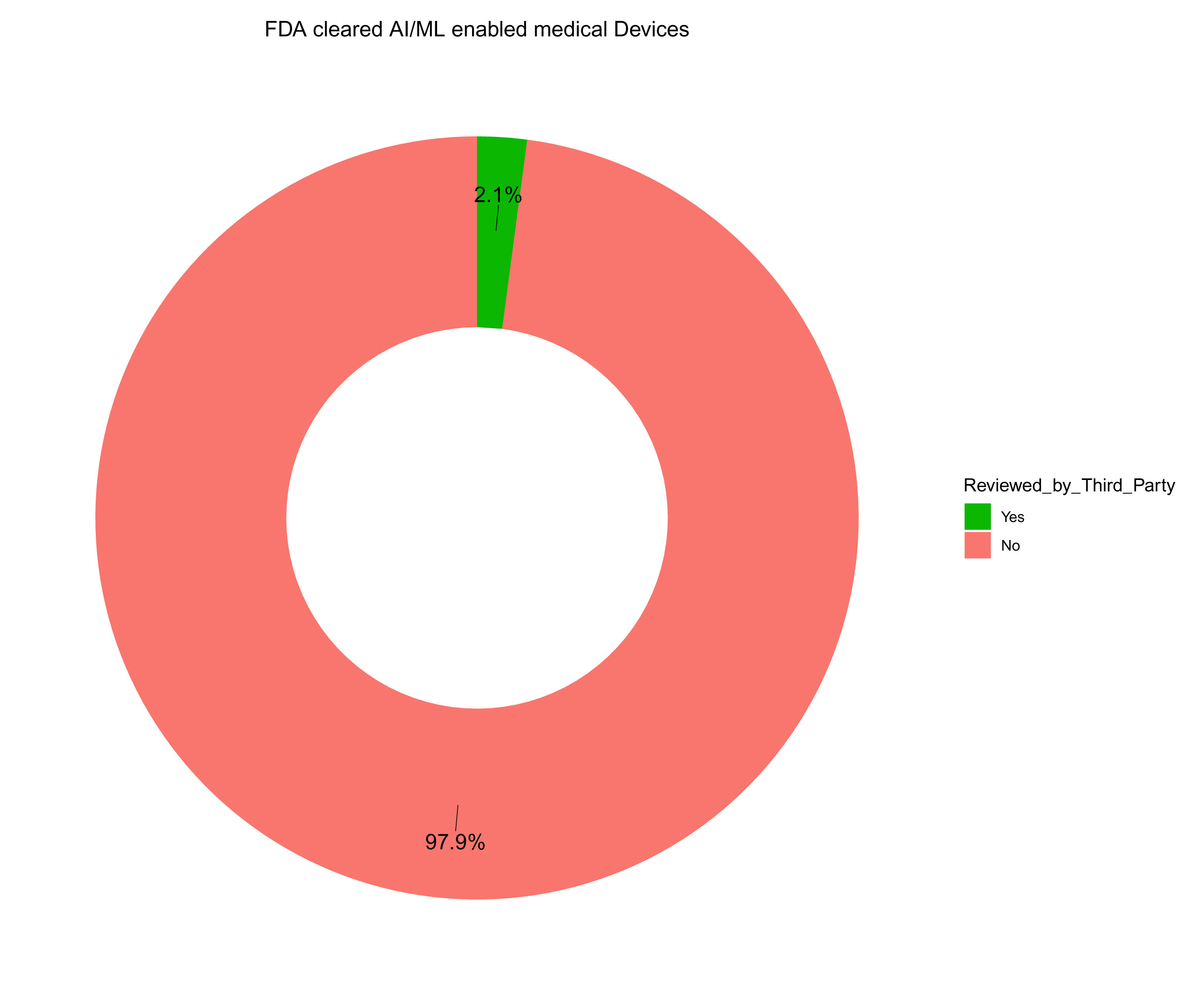

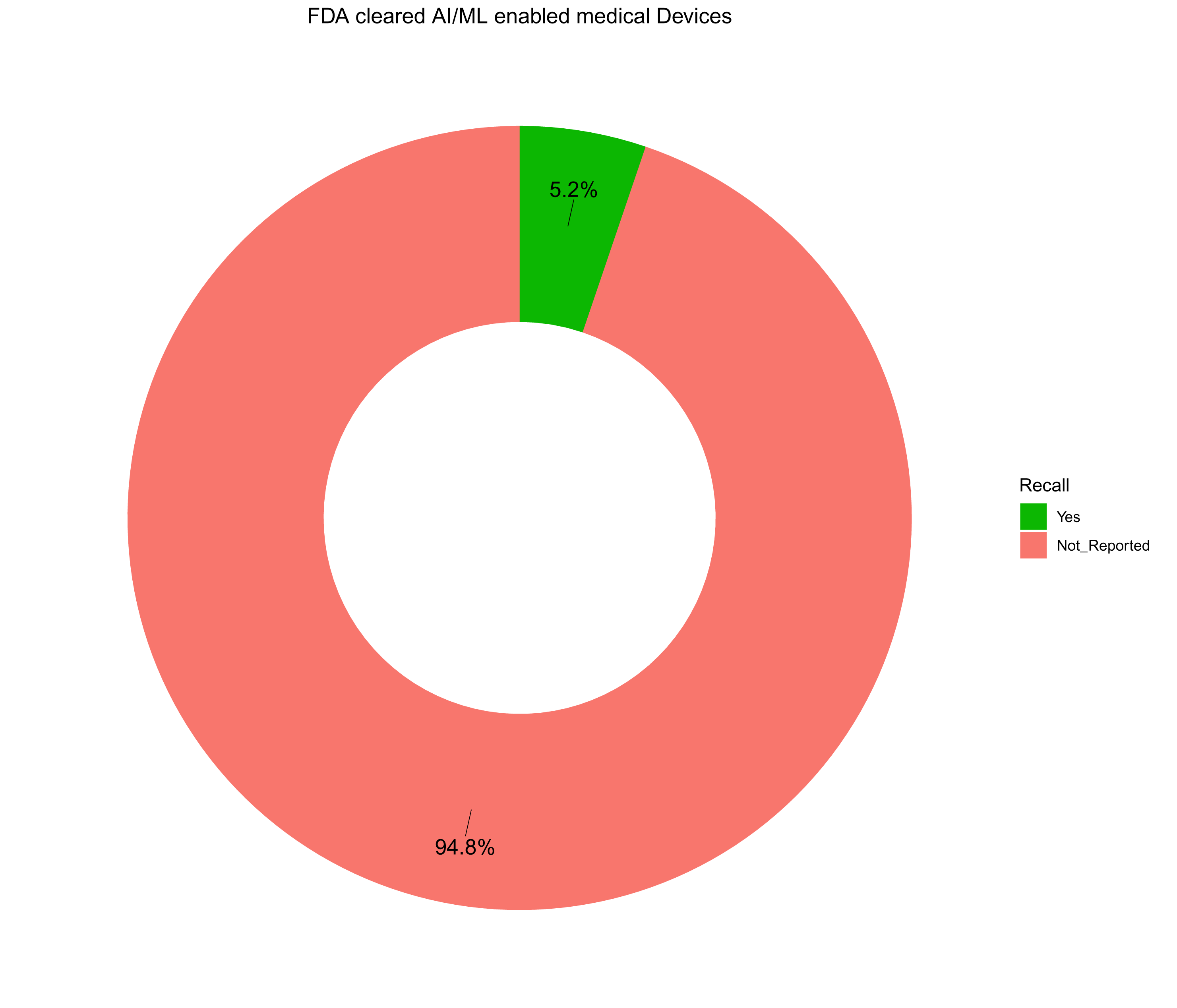

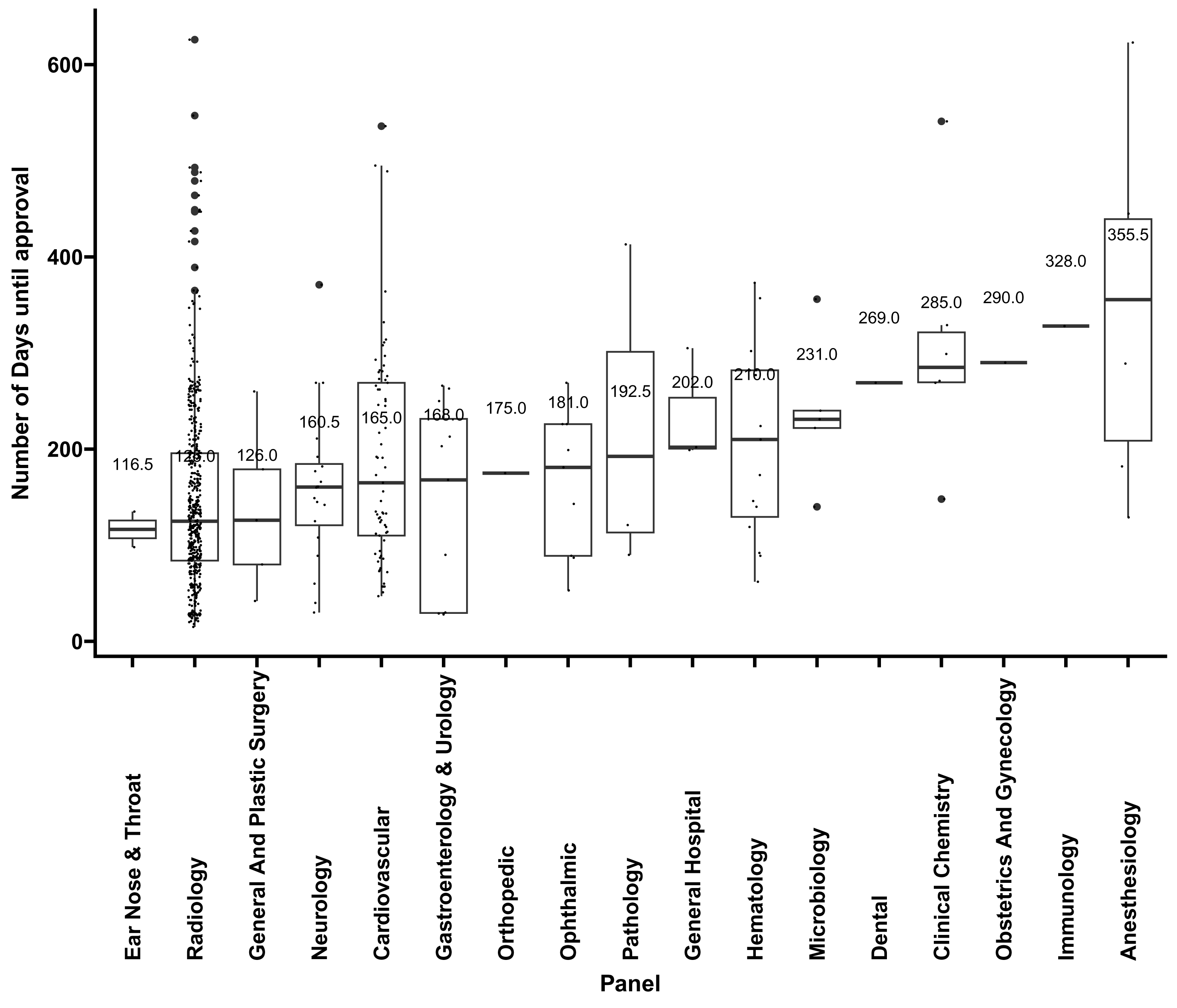

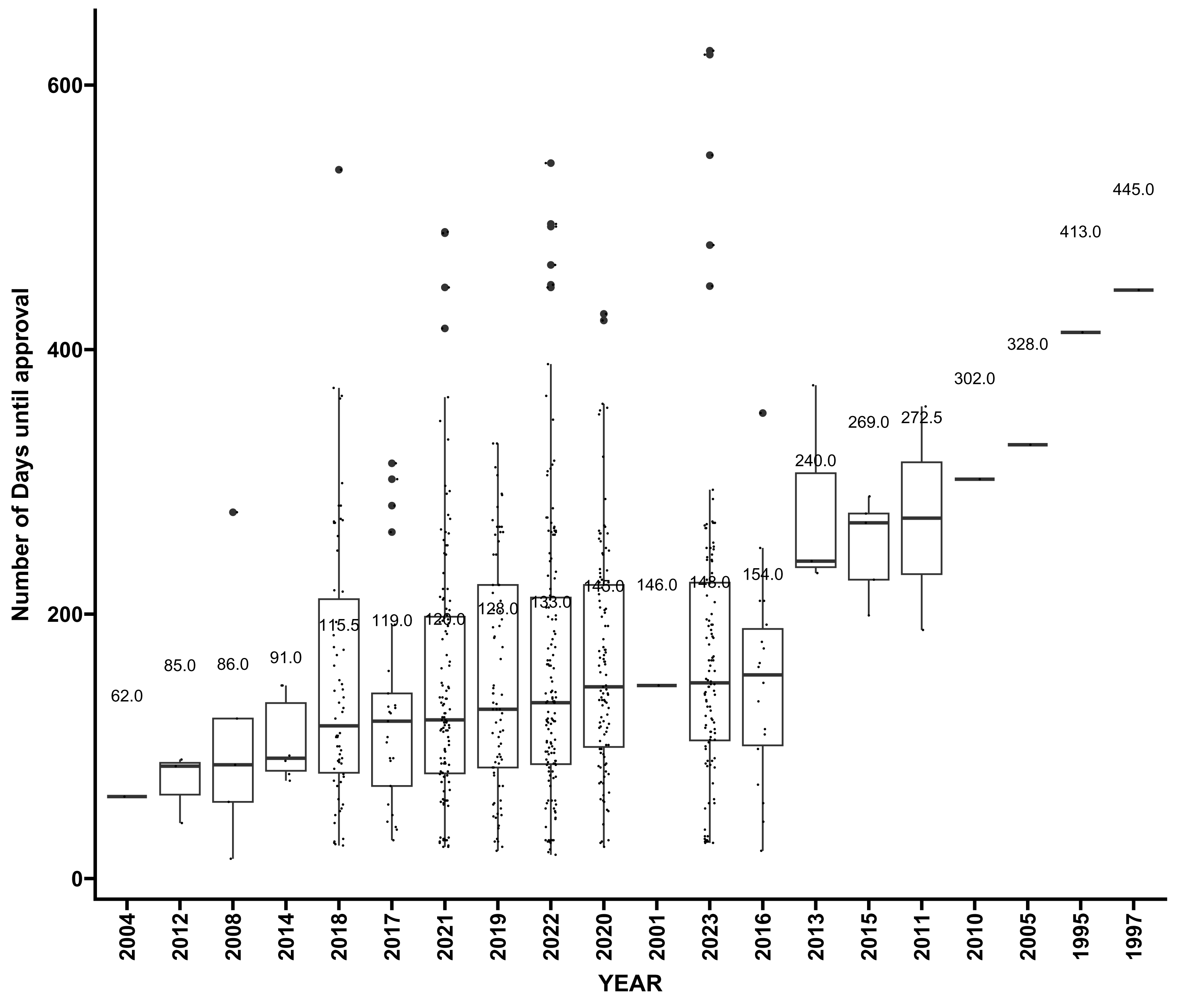

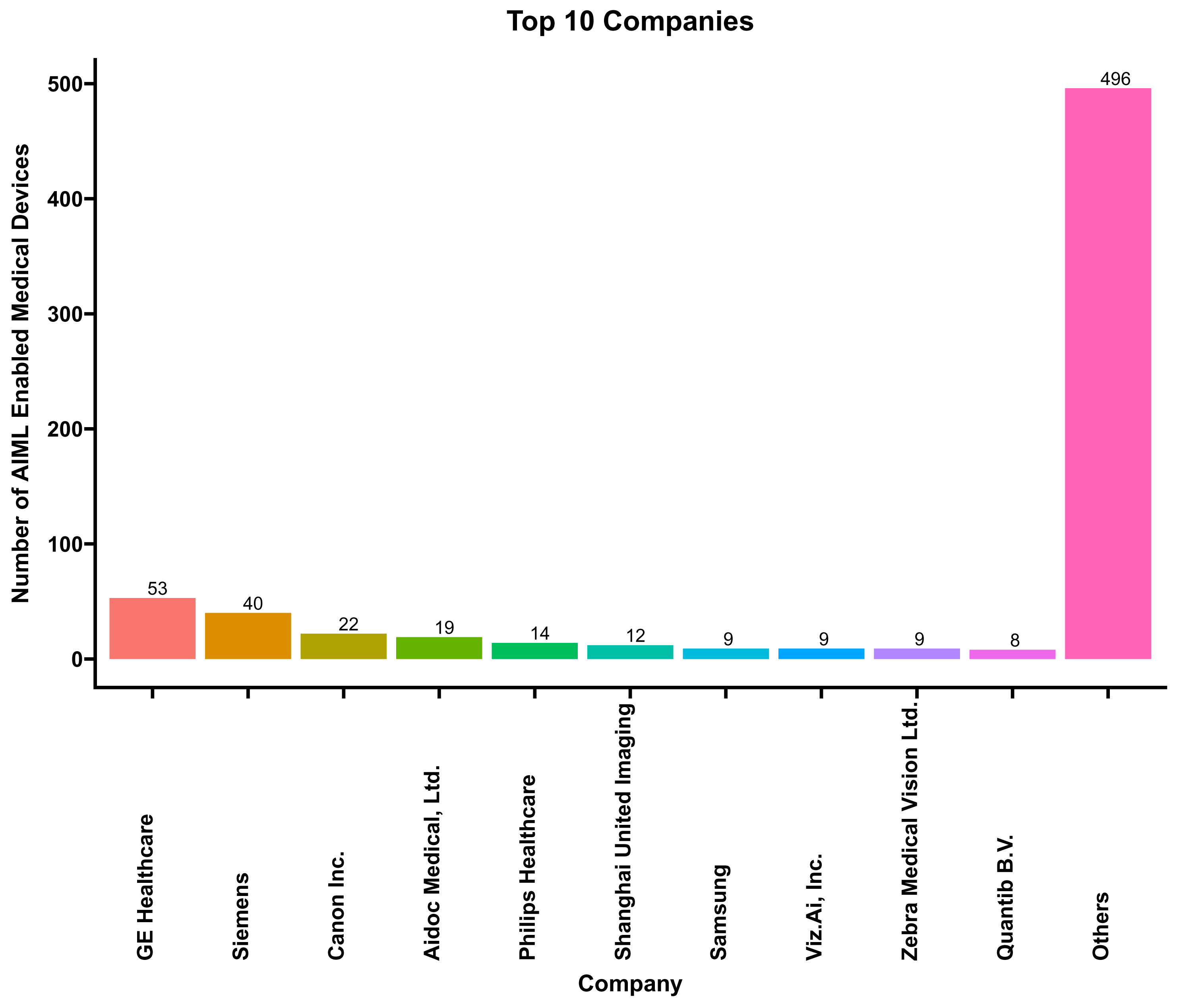

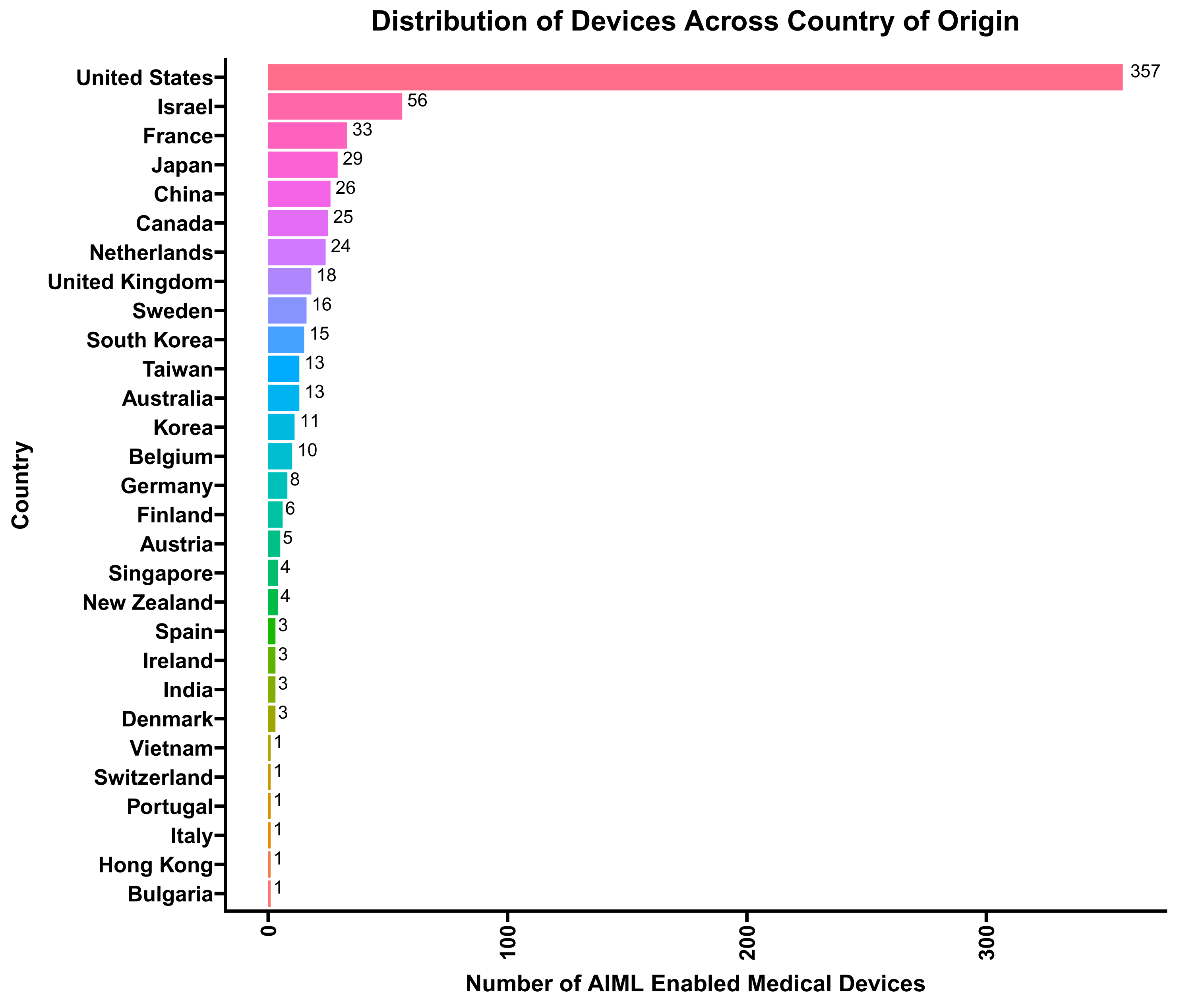

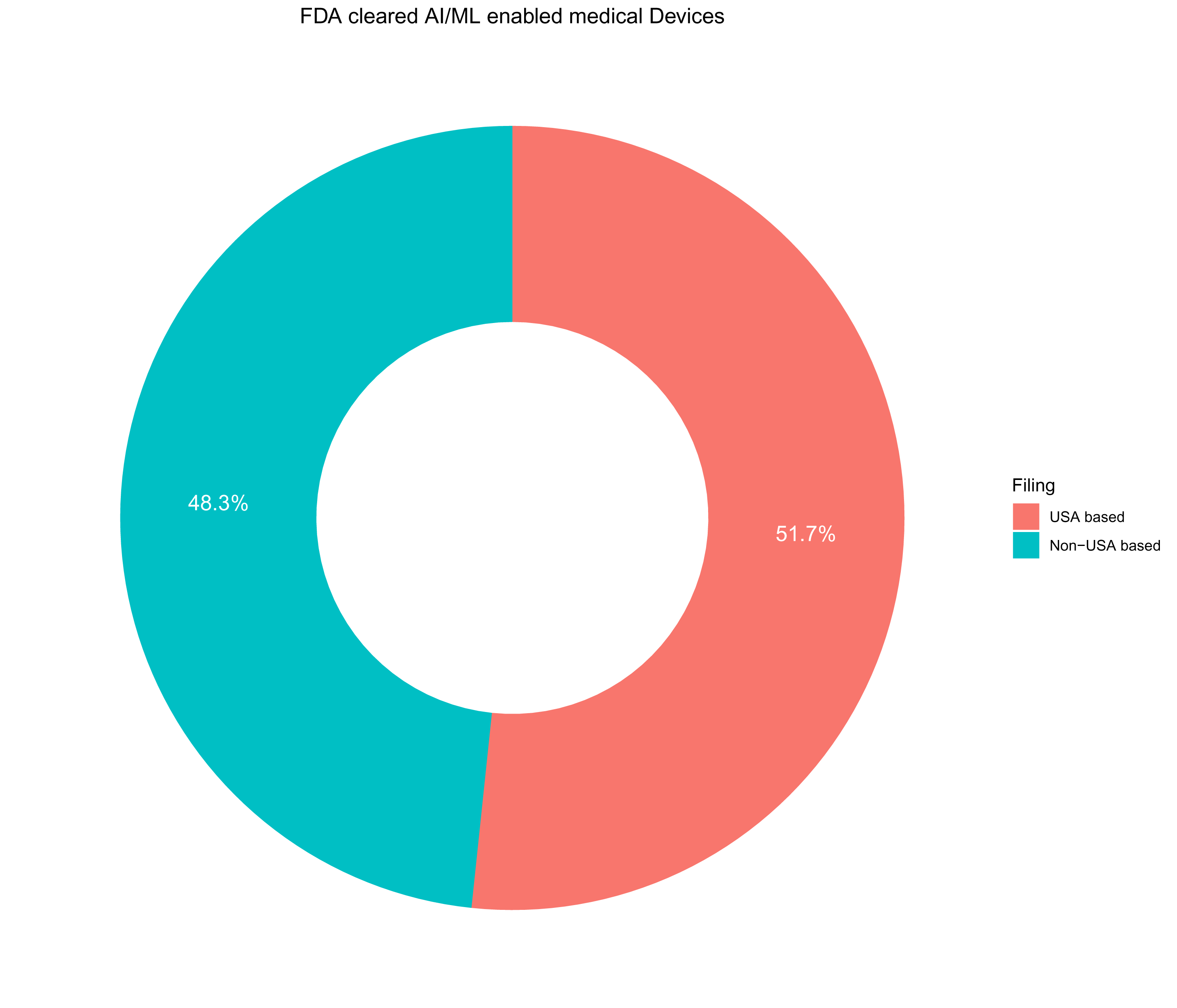

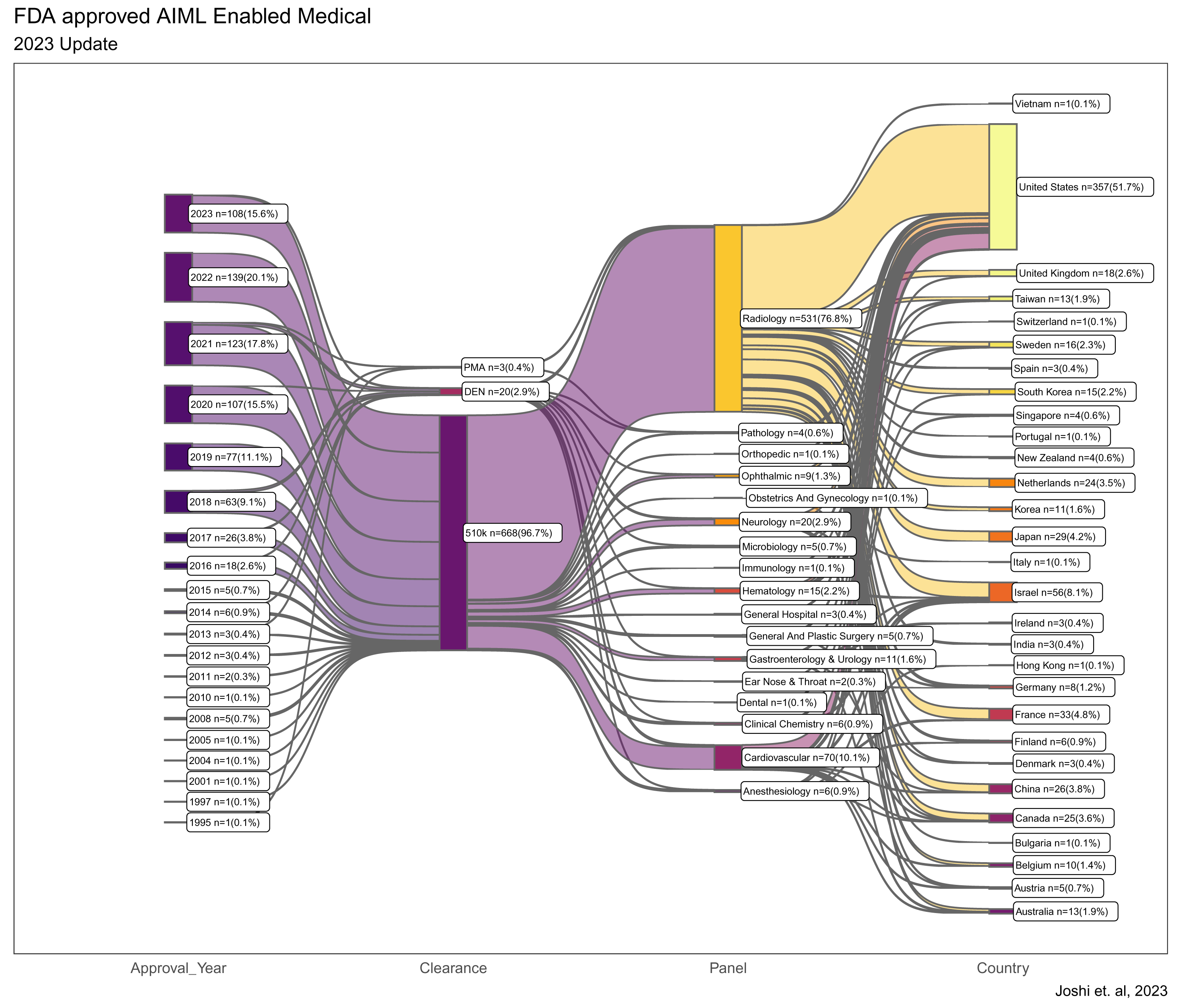

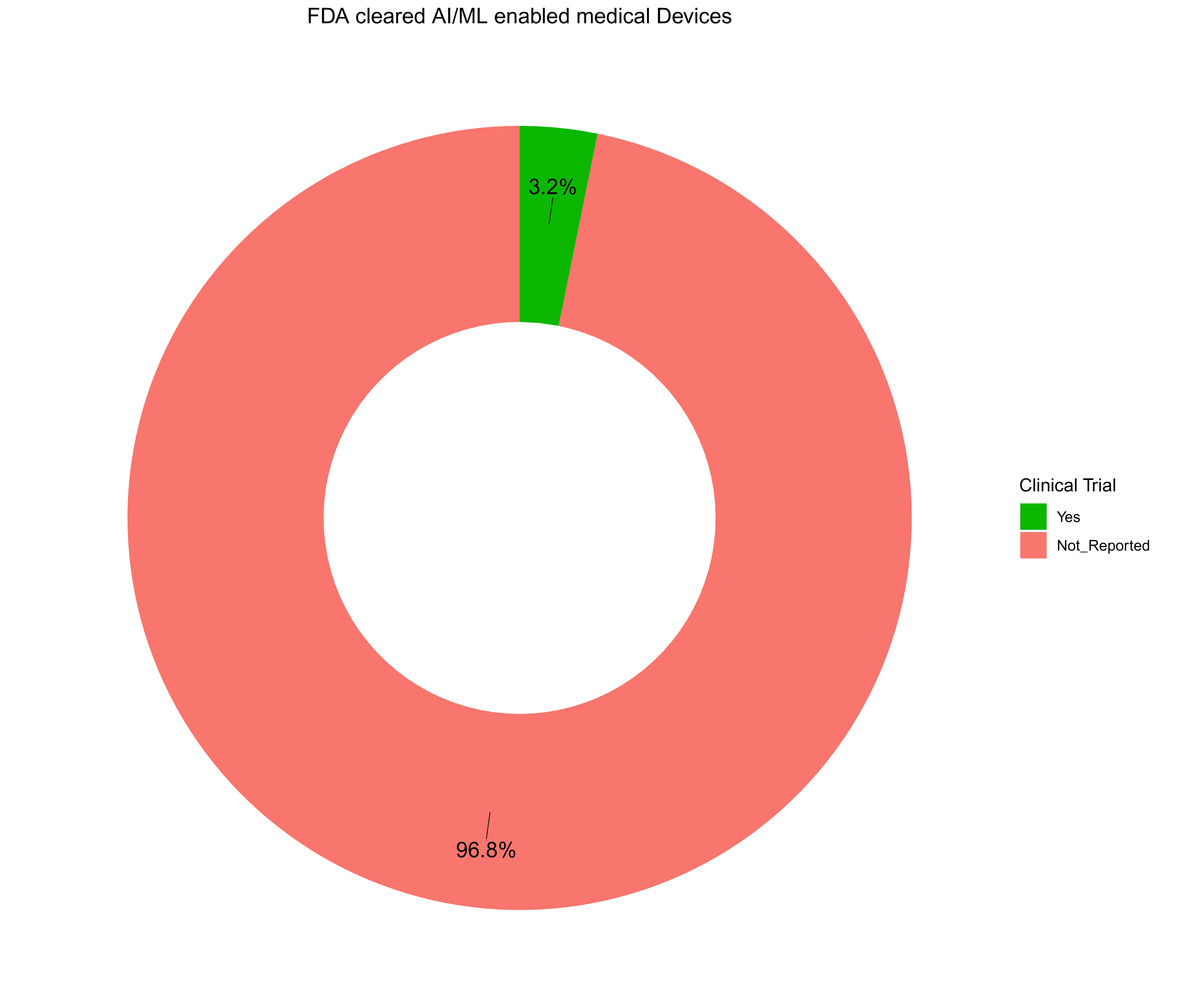

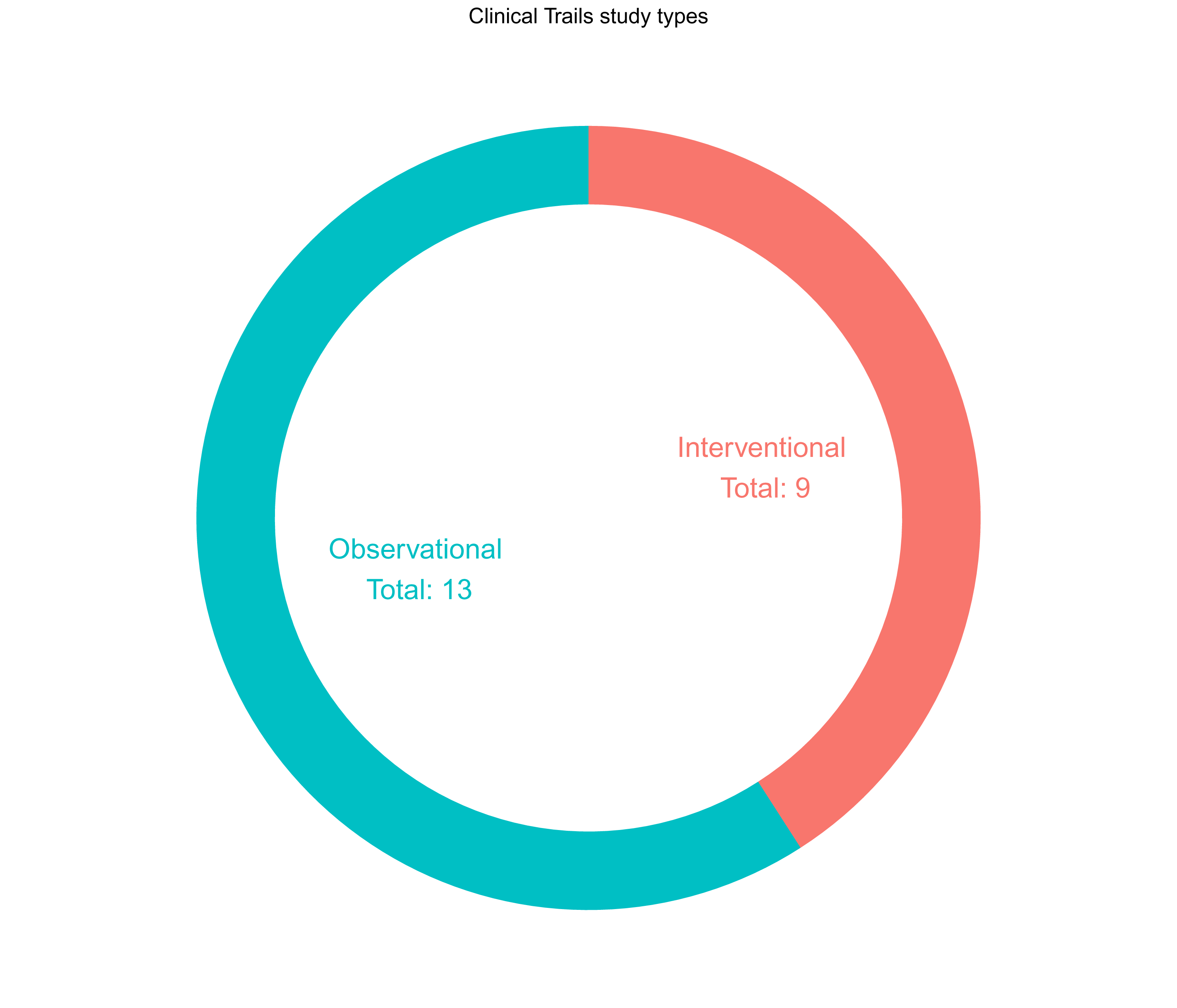

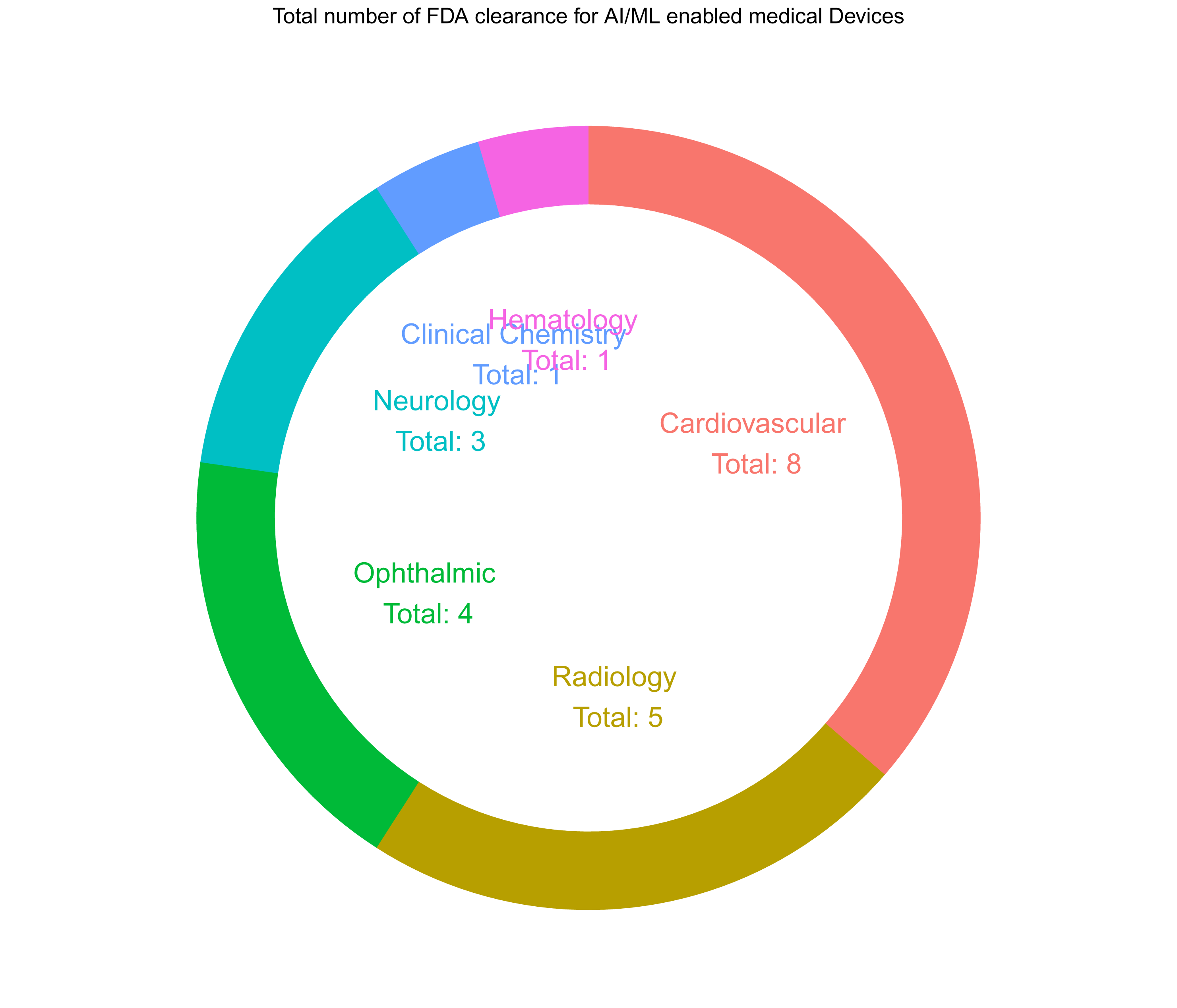

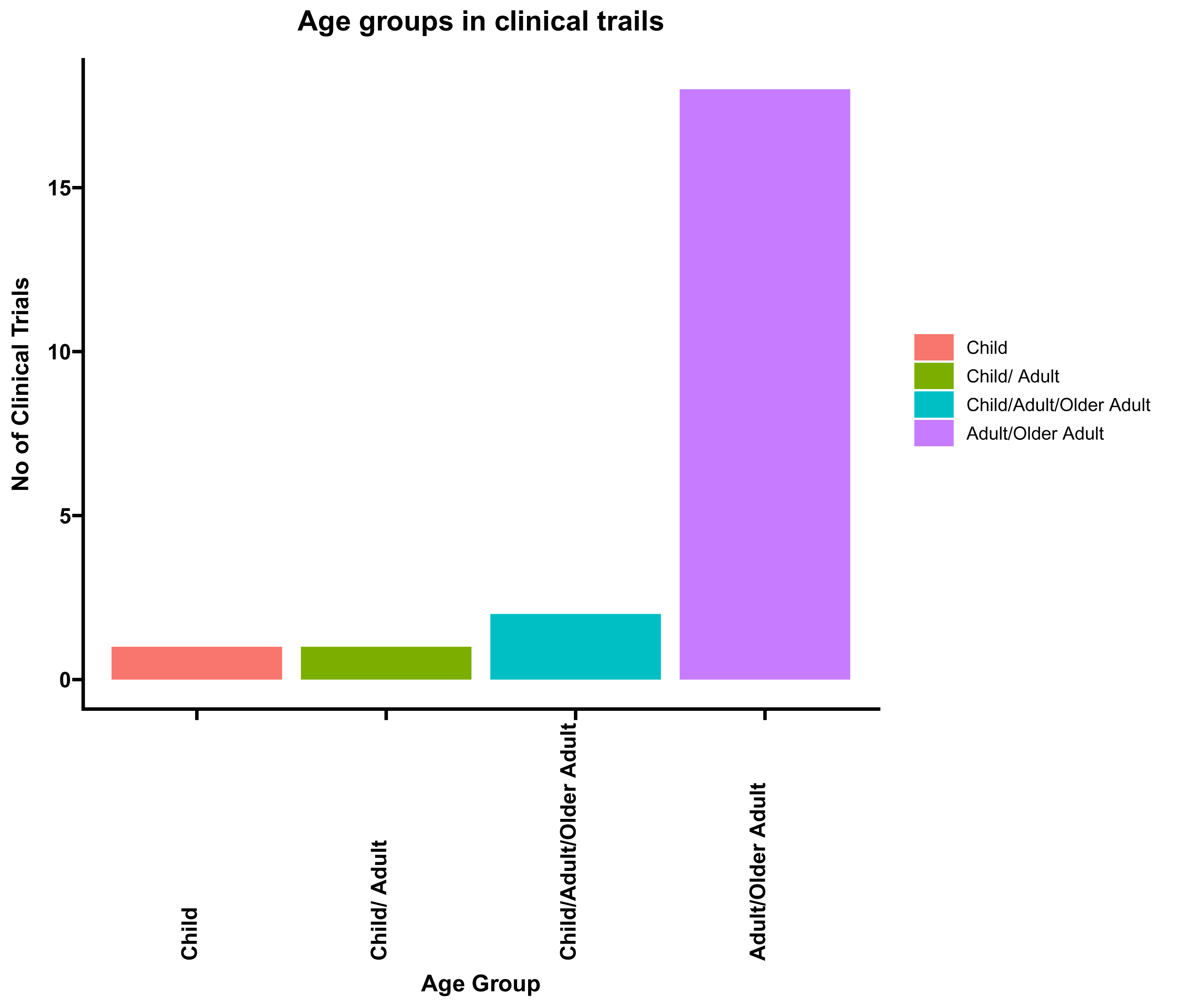

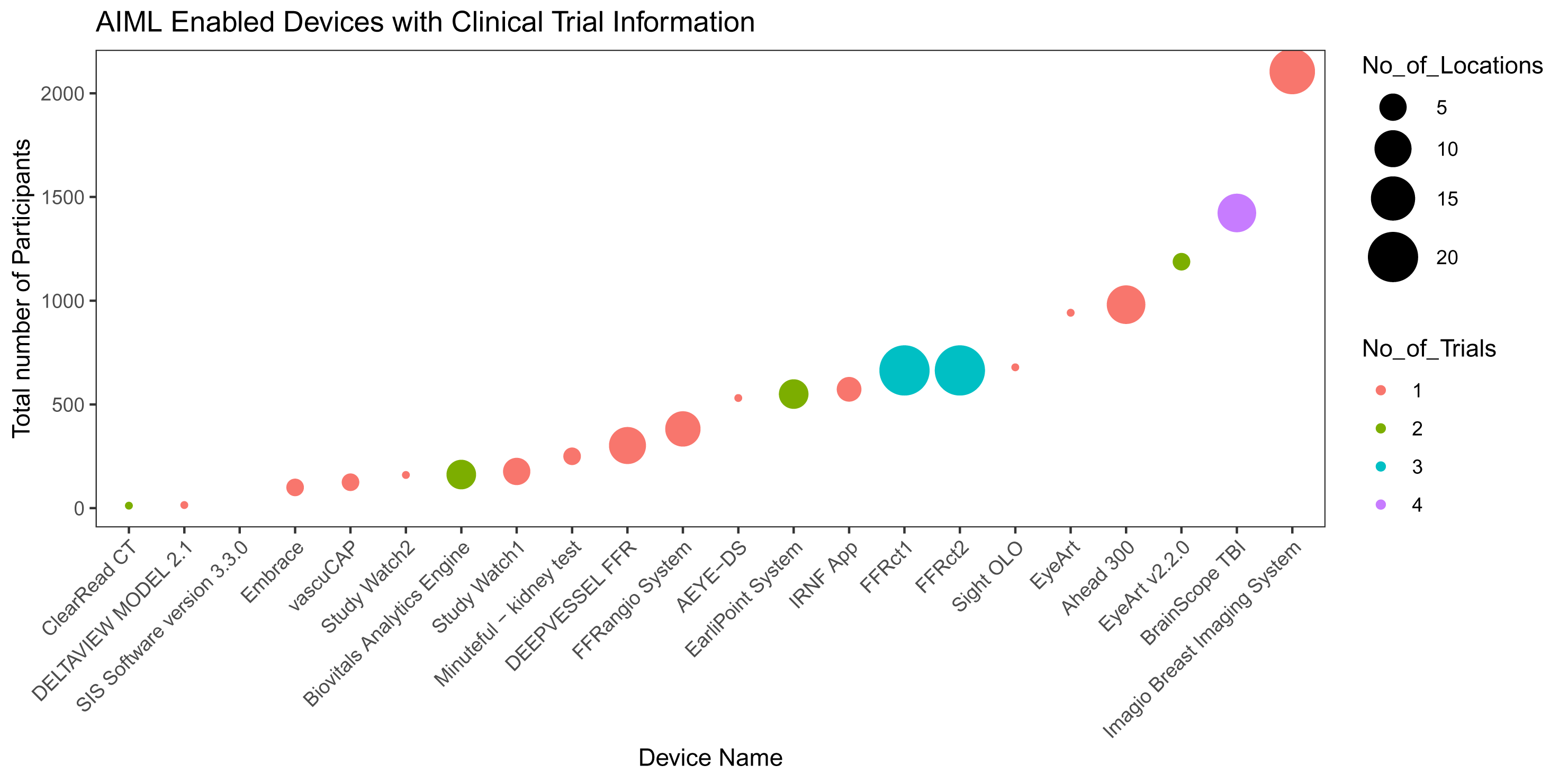

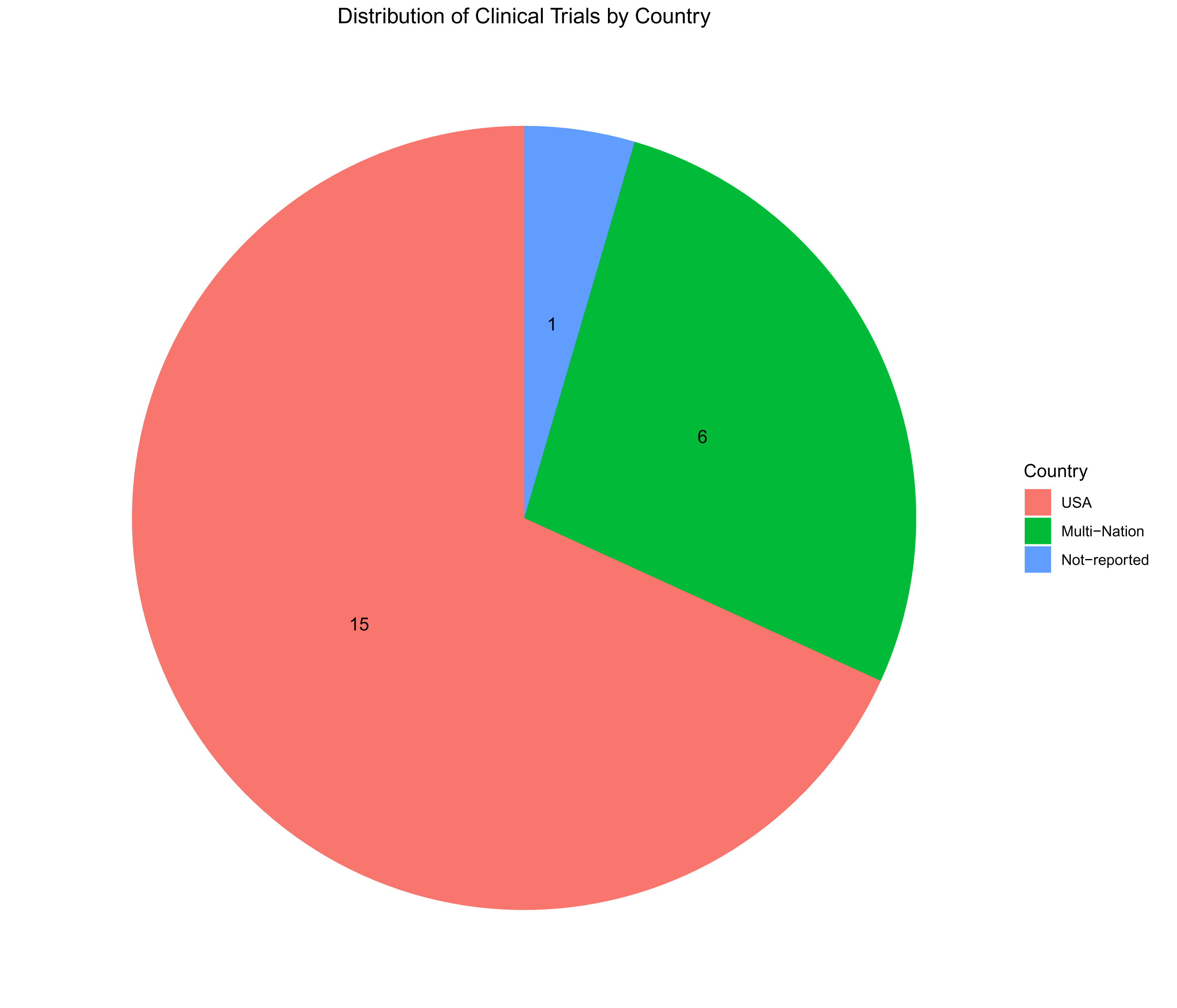

