## Supplemental Table for "FDA approved Artificial Intelligence and Machine Learning (AI/ML)-Enabled Medical Devices: An updated landscape": Supplementary_tables.docx

**Supplementary Table 1: FDA approval pathways and sub-pathways for AI/ML-based devices**

| **FDA regulatory Description pathway** | **Description** |
| --- | --- |
| 510(k) pathway | **Traditional 510(k):** depends on demonstration of Substantial Equivalence **Abbreviated 510(k):** depends on the use of guide documents, controls and recognized standards  **Special 510(k):** when device modifications are made to a manufacturer's own legally marketed device |
| De Novo | **Option 1:** After obtaining a high-level not substantially equivalent (NSE) determination **Option 2:** After determining that there is no legally marketed device |
| Premarket approval (PMA) | **Traditional PMA:** include application with device description and intended use, nonclinical/clinical studies, case report forms, manufacturing methods, labeling **Modular PMA:** for medical products in early stages of clinical study  **Product Development Protocol:** for devices that uses well established technology and serves as a contract upon details of design and development activities **Humanitarian Use Device (HUD): for** device intended to benefit not more than 8,000 patients in the treatment or diagnosis of a disease or condition |

**Supplementary Table 2: Primary product codes and related device classification**

| Primary Product Code | Device Classification Name |
| --- | --- |
| IYN | system, imaging, pulsed doppler, ultrasonic |
| JAK | system, x-ray, tomography, computed |
| LLZ | system, image processing, radiological |
| LNH | system, nuclear magnetic resonance imaging |
| MUJ | system, planning, radiation therapy treatment |
| QAS | radiological computer-assisted triage and notification software |
| QDQ | radiological computer assisted detection/diagnosis software for lesions suspicious for cancer |
| QFM | radiological computer-assisted prioritization software for lesions |
| QIH | automated radiological image processing software |
| QKB | radiological image processing software for radiation therapy |
